# Regulating Flexibility for Artificial Intelligence FDA Experience with Predetermined Change Control Plans

**DOI:** 10.1101/2025.08.26.25334477

**Authors:** Kyra L. Rosen, Kenneth D. Mandl

## Abstract

**Importance:** Predetermined Change Control Plans (PCCPs) are a recent regulatory innovation by the U.S. Food and Drug Administration (FDA) introduced to enable dynamic oversight of artificial intelligence and machine learning (AI/ML)-enabled medical devices.

**Objective:** To characterize FDA program of PCCPs among AI/ML-enabled medical devices, including device characteristics, preapproval testing, planned modifications, and post-clearance update mechanisms.

**Design:** This cross-sectional study reviewed FDA-cleared or approved AI/ML-enabled medical devices with authorized PCCPs.

**Setting:** AI/ML-enabled devices approved or cleared prior to May 30, 2025 were identified from an FDA-maintained public list and their characteristics extracted from FDA approval databases.

**Participants:** N/A

**Main Outcome(s) and Measure(s):** Primary outcomes included (1) prevalence and characteristics of devices with authorized PCCPs, (2) types of FDA-authorized modifications, (3) presence and nature of preapproval testing, such as study design and subgroup testing, and (4) postmarket device update mechanisms and transparency.

**Results:** Among 26 identified AI/ML-enabled medical devices with authorized PCCPs, 92% were cleared via the 510(k) pathway, and all were classified as moderate risk. Devices were primarily intended for use in diagnosis or clinical assessment, and six had consumer-facing components. Authorized modifications spanned the product lifecycle, most commonly allowing model retraining (69% of devices), logic updates (42% of devices), and expansion of input sources (35% of devices). Preapproval testing was limited with seven devices prospectively evaluated and thirteen undergoing human factors testing. Subgroup analyses were reported for eleven devices and none included patient outcomes data. No postmarket studies or recalls were identified. User manuals could be identified online for 54% of devices, though many lacked performance details or mentioned PCCPs.

**Conclusions and Relevance:** FDA authorization of PCCPs grants manufacturers substantial flexibility to modify AI/ML-enabled devices postmarket, while preapproval testing and postmarket transparency are limited. These findings highlight the need for strengthened oversight mechanisms to ensure ongoing safety and effectiveness of rapidly evolving AI/ML-enabled technologies in clinical care.

## INTRODUCTION

The accelerating incorporation of artificial intelligence and machine learning (AI/ML) into medical devices challenges traditional regulatory frameworks, which assume static device characteristics at market authorization.^1-4^ Historically, any substantial modification to a medical device cleared by the U.S. Food and Drug Administration (FDA) required premarket authorization through a resource-intensive new marketing submission.^5^ This process has proven slow and burdensome, making it poorly suited to regulate AI/ML software designed to evolve continuously through retraining or local adaptation.^6-8^

To address this regulatory challenge, the FDA proposed Predetermined Change Control Plans (PCCPs) in 2019, and Congress formally established the FDA’s statutory authority to review and approve them in 2022 by enacting Section 515C of the Food, Drug, and Cosmetic Act through the Food and Drug Omnibus Reform Act (FDORA).^9,10^ PCCPs allow device manufacturers to seek prospective FDA authorization for specified future modifications, bypassing the need for additional submissions. Submitted alongside initial device applications, the FDA recommends that PCCPs detail planned device modifications, methods and standards for repeat safety evaluations, and a plan to communicate changes to existing users. Agency guidance also indicates that authorized modifications should not alter a device’s intended use (general purpose) or, in most cases, indications for use (target disease and patient population). Devices cleared via the 510(k) pathway must also maintain substantial equivalence to their predicate devices.^5^

As PCCPs now represent a central element of the FDA’s regulatory approach to AI/ML-enabled devices, we evaluated their implementation by reviewing all publicly available FDA summaries of AI/ML-enabled medical devices with authorized PCCPs. We also explored PCCP use to date by reviewing publicly available information regarding device updates on device sponsor websites. We sought to characterize how the FDA is operationalizing this adaptive oversight model and to inform future regulatory, clinical, and health-system decision-making.

## METHODS

We included all AI/ML-enabled medical devices with a PCCP authorized by the FDA between November 8, 1995, and May 30, 2025. AI/ML-enabled devices were first identified from a public list maintained by the FDA that includes each device’s name and submission number.^11^ To determine whether each device on the list had an authorized PCCP, we queried FDA regulatory databases by device submission number.^12-14^ The databases cover medical device submission pathways, including 510(k), De Novo, and premarket approval (PMA), and include basic device information such as FDA decision date, applicant, risk classification, and presence of an authorized PCCP. Databases also contain device summaries for all submission types other than PMA supplements — a type of submission used to modify a PMA-approved device — which provide an accessible overview of a device’s risk classification, intended use, technology, performance, and PCCP details. The complete device applications for all approval pathways, including additional testing data and proprietary device features, remain confidential.

Because the FDA’s AI/ML list is described as non-exhaustive, we also queried the same databases for all devices with an authorized PCCP, then reviewed those summaries for terms associated with AI/ML: “artificial intelligence,” “machine learning,” “AI,” “ML,” “deep learning,” “neural net,” and “natural language.” We also searched summaries for a broader computational and software-related terms: “training,” “tuning,” “tune,” “model,” “regression,” “algorithm,” and “software.” While these broader terms do not definitively indicate AI/ML, they identify devices that may raise similar regulatory considerations and, with greater detail in device summaries or broader definitions of AI/ML, may reasonably be classified as such. These devices, because they were not on the FDA-maintained list of AI/ML devices, were excluded from our primary analysis, but were nevertheless reviewed as informative examples of PCCP implementation.

For all identified AI/ML-enabled devices, we extracted data pertaining to device characteristics, preapproval testing, and PCCPs from device summaries. Pediatric was defined as for use in patients <18 years old. Fetal ultrasound devices were classified as for pediatric use based on maternal ages. When summaries explicitly stated that subject and predicate devices were identical or shared preapproval testing, predicate summaries were used for data extraction.

To characterize device monitoring and safety after clearance, we searched public, FDA-maintained databases by submission number for devices in our sample and their predicate devices when the two were identical: Post-Approval Studies (PAS), 522 Postmarket Surveillance Studies, Medical Device Recalls, and Manufacturer and User Facility Device Experience (MAUDE).^15-18^ 522 orders are FDA-mandated postmarket surveillance studies for certain moderate- and high-risk medical devices. The MAUDE database contains medical device adverse outcomes reports, submitted voluntarily by clinicians, patients, or consumers and on a mandatory basis by manufacturers, user facilities (such as hospitals), importers, and distributors.^7,19^

Finally, we estimated the frequency and content of device updates based on public information from device sponsor websites. For each device in our sample, we performed Google searches using device trade name, sponsor name, “instructions for use,” and “user manual.” When sponsor websites were identified, every user manual or instructions for use contained in documentation libraries and/or product pages were examined to match devices in our sample with publicized products. Device name, FDA submission number, and indication for use were used to match devices, given that several devices were named differently on sponsor sites and FDA documents. If relevant documents were identified, we determined whether they included version history, performance metrics, and/or description of the PCCP.

## RESULTS

Twenty-six AI/ML-enabled medical devices with FDA-authorized PCCPs were identified from the FDA list (Table 1). One of them (Caption Guidance, K201992) is a modified version of an earlier approval in our sample — it received 510(k) clearance for a modification to a pre-existing PCCP. We counted the modified version as a distinct device because it relied on updated preapproval testing and reported changes to the PCCP. As of September 2024, 1.9% of AI/ML-enabled devices had authorized PCCPs.

**Table 1:** Characteristics of AI/ML-Enabled Devices with Authorized PCCPs.

| **Device Name (Submission Number)** | **Sponsor** | **Date of Approval** | **Intended Use** | **Primary User** | **Use Population Age** |
| --- | --- | --- | --- | --- | --- |
| Caption Guidance (DEN190040) | Bay Labs, Inc. | 2/7/20 | Echocardiogram acquisition | Provider | Adult |
| Caption Interpretation Automated Ejection Fraction Software* (DEN220063) | Caption Health, Inc | 2/24/23 | Echocardiogram interpretation | Provider | Adult |
| Caption Guidance* (K201992) | Caption Health | 9/18/20 | Echocardiogram acquisition | Provider | Adult |
| LINQ II Insertable Cardiac Monitor, Zelda AI ECG Classification System (K210484) | Medtronic, Inc. | 6/11/21 | ECG interpretation | Patient and provider | Adult |
| Corvair (K231010) | AliveCor, Inc. | 6/7/24 | ECG Interpretation | Provider | Adult |
| Irregular Rhythm Notification Feature* (K231173) | Apple Inc. | 7/21/23 | Arrhythmia detection from pulse rate | Patient | Adult |
| REMI AI Discrete Detection Module (K231779) | Epitel, Inc. | 1/3/24 | EEG interpretation | Provider | Adult and pediatric |
| Low Ejection Fraction AI-ECG Algorithm (K232699) | Anumana, Inc. | 9/28/23 | ECG interpretation | Provider | Adult |
| BoneMRI (K233030) | MRIGuidance B.V | 3/1/24 | MRI processing | Provider | Adult and pediatric |
| CLEWICU System (K233216) | Clew Medical Ltd. | 1/13/24 | EMR-integrated risk prediction | Provider | Adult |
| SleepStageML (K233438) | Beacon Biosignals, Inc. | 3/8/24 | Polysomnography interpretation | Provider | Adult |
| Clarius OB AI (K233955) | Clarius Mobile Health Corp. | 6/14/24 | Ultrasound interpretation | Provider | Adult |
| Acorn 3D Software; Acorn 3DP Model (K234009) | Mighty Oak Medical | 7/12/24 | CT and CTA processing and 3D printing | Provider | Not described |
| AiMIFY (1.x) (K240290) | Subtle Medical, Inc. | 8/21/24 | MRI processing | Provider | Adult and pediatric |
| Tyto Insights for Crackles Detection (K240555) | Tyto Care Ltd. | 7/2/24 | Stethoscope-integrated audio interpretation | Patient and provider | Adult and pediatric |
| Sleep Apnea Notification Feature (K240929) | Apple Inc. | 9/13/24 | Sleep apnea detection from breathing pattern data | Patient | Adult |
| AI Platform 2.0 (AIP002) (K240929) | Exo Imaging | 8/5/24 | Ultrasound acquisition and processing | Provider | Adult |
| FETOLY-HEART (K241380) | Diagnoly | 9/11/24 | Ultrasound acquisition | Provider | Not described |
| Overjet Charting Assist* (K241684) | Overjet, Inc | 8/27/24 | Automated charting | Provider | Adult and pediatric |
| syngo Dynamics (Version VA41D) (K242551) | Siemens Healthcare GmbH | 4/3/25 | Multimodal image processing and interpretation | Provider | Adult and pediatric |
| HeartFocus (V.1.1.1) (K242807) | Deski | 4/4/25 | Ultrasound acquisition | Provider | Adult |
| Tyto Insights for Rhonchi Detection (K243567) | Tyto Care Ltd. | 4/7/25 | Stethoscope-integrated audio interpretation | Patient and provider | Adult and pediatric |
| Canvas Dx* (K243558) | Cognoa Inc. | 4/11/25 | Autism Spectrum Disorder diagnostic aid | Patient and provider | Pediatric |
| Clarius Prostate AI (K243853) | Clarius Mobile Health Corp. | 4/16/25 | Ultrasound processing | Provider | Adult |
| BrightHeart View Classifier (K243684) | BrightHeart | 5/7/25 | Ultrasound acquisition | Provider | Adult |
| Clarius Median Nerve AI (K250226) | Clarius Mobile Health Corp. | 5/8/25 | Ultrasound processing | Provider | Adult |

\* = Device is identical to its predicate other than the addition of a PCCP

FDA database queries identified 74 device applications with authorized PCCPs. Forty-two were cleared via the 510(k) pathway, two via the De Novo pathway, and 30 were PMA supplement approvals. All identified AI/ML-enabled devices were already present on the FDA list. However, four devices on the FDA’s list were not retrieved through database queries, despite discussing authorized PCCPs in their summaries. Among all devices with authorized PCCPs identified through database queries, 29.7% were AI/ML-enabled and 45.9% contained software or computational algorithm components.

Twenty-four AI/ML-enabled devices with PCCPs were cleared through the 510(k) pathway and two were granted marketing authorization through the De Novo pathway. All identified devices received class II, or moderate risk, designations. One (LINQ II) is implantable. Sixteen are approved for use in adults only, seven for adults and children, and one for children only. Six were approved prior to 2024, thirteen in 2024, and seven in the first five months of 2025.

The most common intended uses for these devices are signal interpretation, processing, and acquisition (Table 1). All but one device (Overjet) assist in diagnosis or clinical assessment. Six devices have a consumer-facing component, four of which require physician interpretation. Two are mobile health monitoring devices, both sponsored by Apple Inc., and two are remote diagnostic aids, both sponsored by Tyto Care Ltd. Five devices sought FDA approval solely for the authorization of a PCCP – these devices are otherwise identical to their predicates.

### Predetermined Change Control Plans

Two device summaries contained no data regarding authorized PCCPs; therefore, no information could be extracted.

FDA-authorized device modifications are summarized in Figure 1 and detailed in Supplemental Table 1. They span multiple stages of the lifecycle, including preprocessing changes (e.g. audio signal filtering), expansion of indications for use (e.g. to new diseases or age groups), and postprocessing features (e.g. updated output display). The most common authorized change, permitted for 18 devices, was retraining with new data, often to improve performance. Other frequently granted modifications include changes to model architecture (n=11), such as parameter adjustments, and validation of new input sources without changing input type (n=9), including compatibility with additional ultrasound probes or MRI scanners. Summaries provided insufficient detail about modification testing procedures for us to meaningfully evaluate study designs. Sponsors of four devices (Overjet, Tyto for Crackles, Tyto for Rhonchi, Canvas Dx) plan to use real-world device use data for retraining or testing of retrained models and two specify other data sources (Corvair and HeartFocus); the remaining twenty PCCPs do not specify data sources for retraining or modification. Seven PCCPs state that data used for modification testing will be from diverse populations or populations representative of the intended users. One summary describes plans to evaluate modification performance by demographic subgroups (HeartFocus).

**Figure 1.**
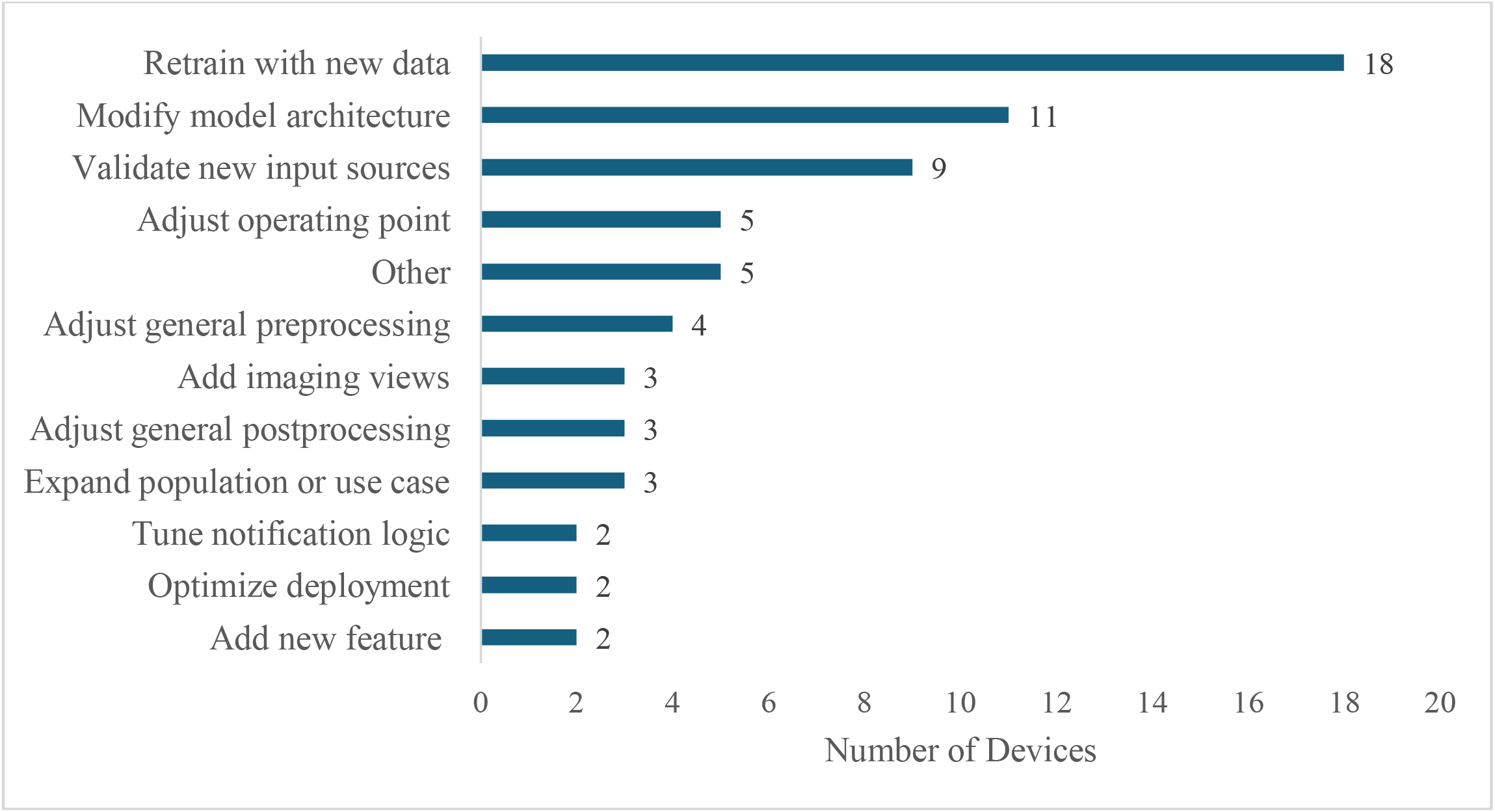
Types of FDA-Approved Modifications for AI/ML Devices

No PCCPs include provisions for adaptive learning, in which devices autonomously update without human oversight. One PCCP (CLEWICU, an ICU-based clinical decision-support tool) includes plans for local tuning to optimize model performance based on local data types and availability but does not specify whether tuning will occur at individual hospitals or across groups of hospitals.

Sixteen PCCPs include methods to update device users when authorized modifications are implemented. Thirteen describe plans to proactively notify users of updates and ten explicitly state that updated performance metrics will be made available.

### Pre-clearance testing

All device summaries include descriptions of pre-clearance testing (Supplemental Table 2).

Seven devices underwent prospective evaluation with a median sample size of 240 participants (IQR 145-499). For one of these devices evaluated prospectively (AI Platform 2.0), sample size was reported as the number of ultrasound examinations obtained. Eighteen devices underwent retrospective evaluation, with a median sample size of 193 patients (IQR 126-445). One retrospectively evaluated device reported sample size as number of images (Overjet), one did not specify the unit of sample size (syngo), and one did not report sample size (Corvair).

Eighteen device summaries describe multisite performance testing; the rest did not report the number of sites. No summaries describes patient health outcomes data.

Eighteen summaries mention the demographics of performance testing samples, of which ten report the distribution of age, eleven of sex/gender, four of BMI, and four of race/ethnicity (Figure 2). Eleven summaries mention subgroup analyses within performance testing, of which four report results by age, two by sex/gender, two by BMI, and one by race/ethnicity (Figure 2). One summary notes that full subgroup analysis results are described in the device label (Caption Interpretation Automated Ejection Fraction Software).

**Figure 2.**
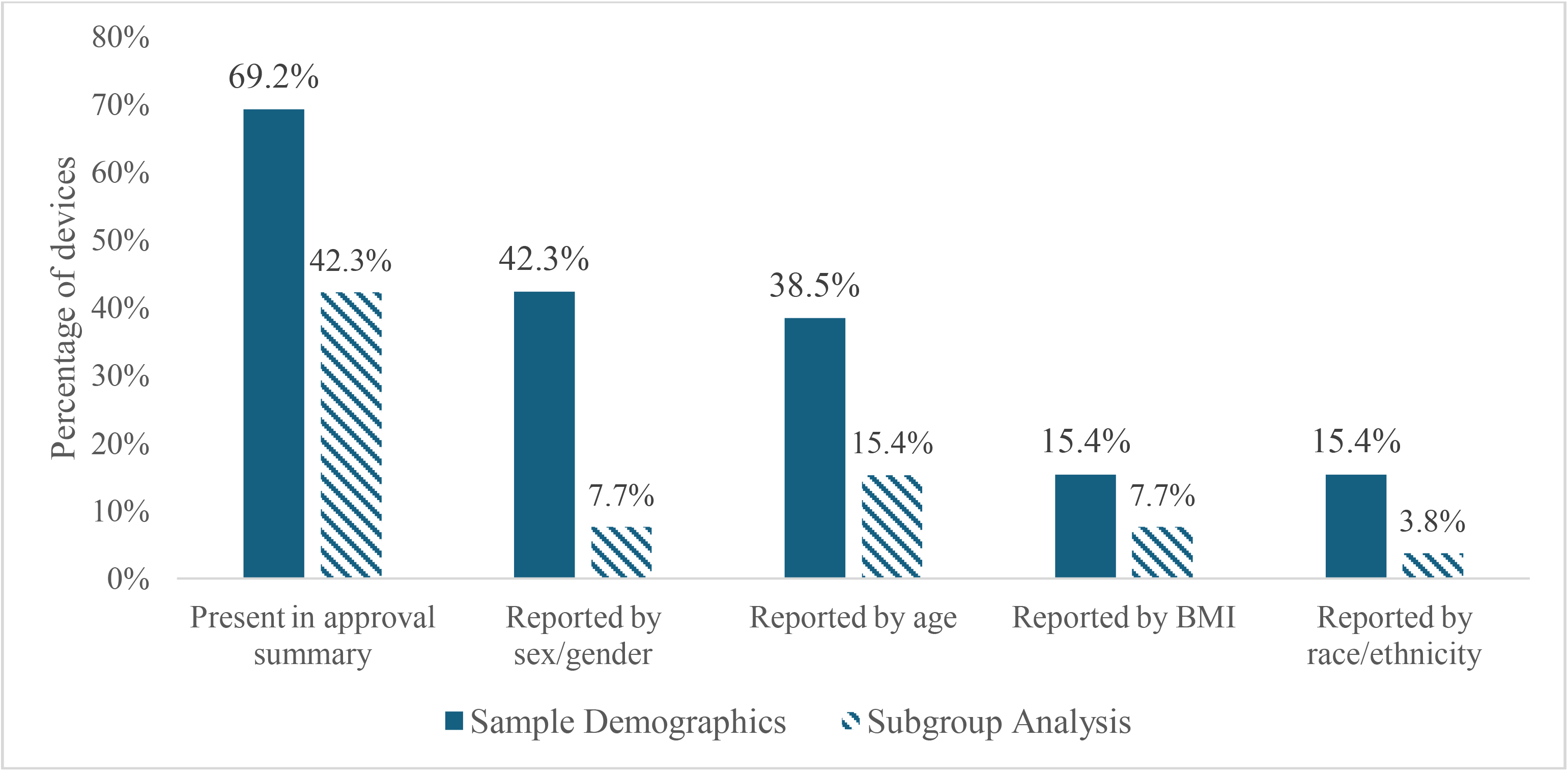
Reporting of Demographic Data and Subgroup Analyses in Preapproval Testing

Thirteen summaries describe human factors testing (Supplemental Table 3). Language in two summaries suggests human factors testing but lacked sufficient detail for confirmation (Acorn 3D, SleepStageML). Three of these summaries report sample sizes, with a median of 31 device users. Of the eleven summaries that report which metrics were evaluated during human factors testing, eight examined task completion or device usability, two measured harmful events, and four assessed device impact on user decision making.

### Postmarket monitoring

One cleared device with a PCCP (LINQ II) has eight safety events reported in the FDA’s Manufacturer and User Facility Device Experience Database (MAUDE). All events were classified as program or algorithm execution problems involving incorrect arrhythmia classification.

Following manufacturer review, four events were attributed to user confusion rather than device error, one lacked sufficient data for assessment, and one was a true error. Two events remain under review. No patient complications were reported.

The FDA has not reported a 522, PAS, or recall order for any AI/ML-enabled device with an authorized PCCP or for identical predicates.

### Sponsor-published materials

User manuals and/or instructions for use (IFUs) were identified on sponsor websites for fourteen devices. Device names differed between FDA documents and user manuals or IFUs for five devices. Twelve user manuals or IFUs describe quantitative performance metrics and eight mention the presence of an authorized PCCP.

Two devices in our sample may have implemented PCCP-authorized updates (Caption Guidance and Corvair). Corvair is described as Kardia12L in its IFU. Three IFU versions are available on the sponsor website, but we were unable to identify differences between them. It is thereby unclear whether the device was modified in line with its authorized PCCP.

Caption Guidance received an original De Novo approval followed by a 510(k) clearance to authorize modifications to its pre-existing PCCP. These were counted as two devices in our sample to account for differences in PCCP and testing characteristics; however, one user manual applies. Caption Guidance is described as Caption AI in the user manual and is integrated into two different Caption devices. Both devices have published version histories with brief descriptions of modifications that are non-identical. It is unclear whether this device’s PCCP was relied upon to authorize these changes.

## DISCUSSION

Use of PCCPs in AI/ML-enabled device clearances is accelerating, with most authorized devices classified as moderate risk, cleared via the 510(k) pathway, and intended for diagnostic or assessment purposes. Approved modifications are broadly defined and span product development and use, providing sponsors with considerable flexibility for postmarket changes. Yet, pre-clearance testing described in summaries was rarely prospective, lacked clinical detail, and had limited use of human factors testing, subgroup analyses, or clinical outcomes. Information on postmarket updates and testing was similarly limited in both FDA and sponsor materials. These findings suggest that, while PCCPs represent a regulatory innovation to accommodate dynamic AI/ML adoption in healthcare, there is a need to strengthen public assurance of device safety and effectiveness before and after modification.

The broad scope of authorized modifications permits substantial changes to device performance and applicability over time, complicating predictions of future safety and effectiveness. Retraining models with new data may improve model performance for certain populations, such as rare or underrepresented cases, but risks degrading performance for others, resulting in edge case failures.^20-22^ Updates to device outputs and postprocessing, such as adding new diagnostic categories and image enhancement features, could keep devices in line with clinical best practice, but may also introduce usability challenges or confusion without additional training and communication.^22,23^ Decision-support devices can reduce physician cognitive load, but silent changes to model parameters or other logic may go unnoticed by habituated users, potentially impairing physician capacity to intervene if performance degrades.^8,24-27^ Ultimately, there is no research into the effects of postapproval device flexibility — whether these modifications will improve or undermine patient safety thereby depends on robust transparency and responsive oversight.^20^

We found that information on pre-clearance evaluation was limited in the device summaries. This aligns with prior work showing a lack of robust premarket evaluation among all AI/ML-enabled medical devices.^1,2,4,28-30^ Limited real-world evaluation of workflow integration and patient outcomes raises particular concern given the risk of performance degradation and the influence of user behavior on device function.^20,22,31-33^ These risks may be amplified as devices are modified. For example, models that are retrained without independent oversight or transparency about the datasets used may be susceptible to covariate shift, degraded external validity, and unanticipated changes in users interaction.^8,20,33,34^ While the majority of AI/ML-enabled devices rely on predicate approvals to supplement safety and efficacy data, they often differ substantially from their predicates in design and function. Even more, predicates themselves frequently rely on other predicate approvals, sometimes across multiple generations.^35^ This compounding of evidence gaps across the development cycles — including PCCPs — risks reducing certainty about device performance.

That being said, a higher proportion of devices with PCCPs, compared to all AI/ML devices, were prospectively evaluated, report validation results by subgroup, and used data from multiple sites. This may indicate that device sponsors are proactively pursuing more rigorous preapproval testing when seeking PCCP authorization or collaborating more closely with the FDA, given that 510(k) clearance typically does not require robust preapproval testing—clinical data is requested for under 10% of submissions.^7^ While 5% of AI/ML-enabled devices experience recall, no devices in our sample have been recalled so far.^4,35^

Postmarket monitoring is essential for AI/ML-enabled devices with PCCPs, even with robust preapproval testing, because performance in diverse, local environments cannot be comprehensively assessed prior to deployment.^33,36,37^ As a former FDA commissioner noted, “the scale of effort needed” for this monitoring “could be beyond any current regulatory scheme,” and will require joint effort between regulators, device sponsors, and health systems.^6^ To do so, the FDA would require more funding and expanded statutory authority.^38^ Currently, the agency can only require postmarket studies for moderate risk devices under narrow conditions or after an adverse event via 522 orders. This authority is rarely exercised and only 8% of 522-ordered studies were reported as complete as of 2021.^7,39,40^ For postmarket monitoring to be routine and mandatory, this capacity would have to be expanded. Another route is to task the FDA with postmarket surveillance. However, centralized oversight like this would require global adoption of unique device identifiers (UDIs) — codes to track device use in clinical practice — an issue the FDA cannot address because it does not have authority over clinical practice.^8,38,41^ Low UDI adoption additionally stymies health system and collaborative efforts to monitor device implementation. While more funding is needed to explore these options, recent innovations —such as Elsa, a generative AI tool used to streamline the FDA review process — could help reduce resource strain.^42^

Device sponsors and health systems may also play a role in postmarket oversight of PCCP-authorized devices. Sponsors could provide health systems with timely information on deployed modifications to enable local oversight of device performance, perhaps through existing update procedures described in device summaries. Health system leaders, in turn, could advocate for greater transparency through contracting processes. Routine collaboration between sponsors and health systems — such as through shared real-world data— could be beneficial to both parties by supporting iterative development and monitoring. Four devices in our sample already plan to use real-world data for model updating, demonstrating sponsor interest in this data. There are calls for health systems to create local governance structures, establish error reporting pathways, and participate in collaborative data sharing networks.^43-46^

Transparency toward clinicians and patients is equally critical. Presently, device users rely on FDA summaries or sponsor materials—such as user manuals and update notifications—to understand device function. FDA summaries are not updated as devices are modified, and we found them lacking information for informed purchasing or use decisions. Available user manuals contained as much as or less information than FDA summaries, and neither they nor update notifications are FDA reviewed. While more detailed information is likely submitted to the FDA and may appear in device labels, it is not publicly disclosed, limiting utility and trust among physicians and patients.^47,48^ Greater label transparency could promote informed decision making, effective research, and ex post enforcement, as discussed in a recent editorial.^49^ Importantly, transparency must be combined with central stewardship, as neither patients nor clinicians can be expected to parse through complex data on device safety and efficacy.

Finally, it remains unclear how high-risk devices and those incorporating emerging technologies like adaptive learning and generative AI will be overseen by the FDA, and many non-AI/ML devices have already received PCCP authorization. Expansion of PCCPs into these broader areas highlights the need for transparency across all device types.

### Limitations

We relied on publicly available summaries, which omit details present in full FDA submissions. Information was particularly limited for high-risk devices approved via the PMA pathway, limiting our ability to classify these devices as AI/ML-enabled or not. Therefore, we may have excluded AI/ML-devices with authorized PCCPs approved through PMA supplement applications. We may have not been successful locating relevant documents, such as user manuals, due to discrepancies in device or sponsor naming. Additional sponsor-published materials such as device labels and update notifications may contain important information but are not publicly available for review. Third, we were unable to determine whether several higher risk devices approved through the PMA pathway were AI/ML-enabled; these devices likely have different characteristics and testing standards than those included.

## CONCLUSION

The wide latitude permitted in post-clearance device modifications, combined with limited initial device evaluations, minimal publicly available detail on pre-clearance clinical testing, the absence of a publicly accessible database of device labels, and fragmented information on postmarket changes under PCCPs, underscores the need for increased rigor in initial clinical assessments, enhanced information management, and improved postmarket surveillance. Importantly, reducing this fragmentation of device information would better equip clinicians and patients to clearly assess and track device safety, effectiveness, and performance in real-world clinical settings.

## Data Availability

All data presented in the study are publicly available.

## Notes

### Competing Interest Statement

The authors have declared no competing interest.

### Funding Statement

National Center for Advancing Translational Sciences (NCATS), National Institutes of Health (U01TR002623)

