## Supplementary Tables for "Regulating Flexibility for Artificial Intelligence FDA Experience with Predetermined Change Control Plans"

**Supplementary Table 1: Device modifications approved in authorized PCCPs**

| Device Name | Approved Modification(s) |
|---|---|
| Caption Guidance | Not described in approval summary |
| Caption Interpretation Automated Ejection Fraction Software | 1) Training on additional data<br>2) Incorporation of additional 2D TTE views<br>3) Optimization of implementation of core algorithm(s) in software<br>4) Improving algorithm operating speed |
| Caption Guidance | Not described in approval summary |
| LINQ II Insertable Cardiac Monitor, Zelda AI ECG Classification System | 1) Changing the threshold<br>2) Re-training the algorithm on data labeled following the original protocol and data labeled following an alternate protocol<br>3) Pre-training the algorithm |
| Corvair | 1) Improve algorithm performance by retraining with additional data without modifying the architecture |
| Irregular Rhythm Notification Feature | (1) Adjust the numerical threshold at which a tachogram is classified as AF (operating point)<br>(2) While maintaining the same algorithm architecture and number of parameters, retrain algorithm with additional datasets<br>(3) Modification to the number of sequential tachograms that must be classified as irregular in a given time period to surface a notification<br>(4) Modification to the time period of confirmation cycle |
| REMI AI Discrete Detection Module | 1) Expansion of the training data<br>2) Optimization of the algorithm |
| Low Ejection Fraction AI-ECG Algorithm | Not described in approval summary |
| BoneMRI | 1) Re-training to improve ML model performance with additional training data<br>2) Validation of additional scanner support |
| CLEWICU System | 1) CLEW models to be trained and validated for new input data sets following the same validation protocol and meeting the same performance criteria as were used to validate the CLEWICU models described in this 510(k). Due to differences in US hospital electronic medical record systems and critical care patient monitoring protocols, CLEW anticipates that the CLEWICU models may need to be trained for hospitals that present: 1) reduced input data types or reduced frequency of data availability; or 2) additional input data types as compared to the models cleared under this 510(k).<br>2) CLEWICU models to be trained with new datasets, as they become available, to increase the models' sensitivity while maintaining or improving the models' performance specifications |
| SleepStageML | 1) Update of the machine learning model (neural network) to improve sleep staging performance within the intended use population by<br>a. Retraining with an updated training/tuning dataset,<br>b. Retraining with updated hyper-parameters, loss function, optimizer,<br>c. Retraining with updated model selection criteria,<br>d. Retraining with an updated neural network architecture with limitations on model size and type. |

| | |
|---|---|
| | 2) Update of signal preprocessing steps to improve sleep staging performance within the intended use population by updating the parameters of the digital signal processing steps (e.g., filtering) applied to the EEG signals before being input to the machine learning model<br>3) Update probability postprocessing to improve performance within the intended use population by updating the methods by which sleep stages are generated from the model output sleep stage probabilities<br>4) Update of signal quality check to improve performance within the intended use population by updating the criteria/thresholds used to check that the input EEG signals are analyzable |
| Clarius OB AI | 1) Modification to model architecture<br>2) Modification of model training methods and parameters<br>3) Modification of postprocessing algorithms<br>4) Modification of data input sources (Re-training of the OB AI model to expand its use with new data sources) |
| Acorn 3D Software (AC-SEG-4009); Acorn 3DP Model (AC-101-XX) | Not described in approval summary |
| AiMIFY (1.x) | 1) Additional training and/or performance data to:<br>a. Add more gadolinium-based contrast agents<br>b. Add additional patient ages (infants and children),<br>c. Add additional clinical conditions<br>d. Add additional scanner models<br>2) The AiMIFY AI/ML model is planned to be retrained with additional output channels to reduce processing time<br>3) An optional pre-processing feature is planned that consists of an algorithm which will suppress vessel enhancement of the AiMIFY AI/ML model to reduce vessel conspicuity in the AiMIFY-enhanced images and resulting in improved perceived image quality |
| Tyto Insights for Crackles Detection | 1) Modifications related to quantitative measures of performance specifications: Re-training of the ML model with additional data to improve the performance of the re-trained algorithm compared to the original device while the same type and range of input signal is used.<br>2) Modifications related to quantitative measures – technical performance specifications:<br>a. Modification of data preprocessing methodologies<br>b. Data augmentation methodologies<br>c. Architecture and hyper-parameters to improve the performance or the efficiency of the computational resources (running time, memory consumption and CPU utilization).<br>3) Modifications related to device inputs: Expanding the algorithm to include new sources of the same signal type (different model of FDA compatible Stethoscope with equivalent audio acquisition specifications). Modification is limited to expanding to electronic stethoscopes that have FDA 510k clearance (for over-the counter use) at the time that the proposed modification is made. |
| Sleep Apnea Notification Feature (SANF) | 1) Modifications to Breathing Disturbances (BD) computation:<br>a. Adjust the operating point<br>b. Re-train algorithm with additional datasets while maintaining the same algorithm architecture and number of parameters<br>c. Revise signal input module<br>d. Add additional classifier outputs |

| | |
|---|---|
| | e. Modifications to signal quality<br>f. Modifications to post-processing modules.<br>2) Modifications to sleep apnea estimation:<br>a. Modify the number of nightly BD readings required to surface a notification<br>b. Reduce the interval of the notification window<br>c. Modify logic for surfacing a notification based on BDs. |
| AI Platform 2.0 (AIP002) | 1) Modification to architecture, pre/post processing: The AI models in AI Platform 2.0 may be modified in a focused and bound manner, to improve accuracy, efficiency, and adaptability while maintaining the safety and efficacy of the device.<br>2) The training dataset of the AI models may be augmented with a broader and more diverse range of imaging data to enhance model robustness, reduce bias, improve generalizability, and respond effectively to real-world feedback. |
| FETOLY-HEART | 1) Modification of training and/or validation datasets<br>2) Modification of model training hyperparameters<br>3) Heart quality criteria addition/removal |
| Overjet Charting Assist | 1) Improve performance by reducing false positive and false negative outputs based on post-market and real-world data. Specifically, improvements resulting from re-training the ML model with new data. |
| syngo Dynamics (Version VA41D) | 1) Re-training Auto EF MLDSF with additional data<br>2) Optimizations of the Auto EF ML-DSF core algorithms<br>3) Auto EF ML-DSF with expanded input images including contrast support |
| HeartFocus (V.1.1.1) | 1) Retraining of the core algorithms: This modification's scope is the retraining of the AI/ML models in the perspective of validating a new manufacturer prior to an M2. The modification is limited to retraining with additional data without changing the models' architecture or training procedure. It notably describes the data collection to retrain the models and the evaluation plan to validate the performance of the retrained models on the already-cleared ultrasound systems.<br>2) Extend the use of the ML-DSF with new ultrasound systems: This modification's scope is the validation of HeartFocus on new ultrasound systems. It notably describes the data collection and evaluation plan to validate the performance of the AI/ML models on the new ultrasound system.<br>3) Extend the use of the ML-DSF with new operating systems: This modification's scope is the extension of the compatibility with new operating systems. |
| Tyto Insights for Rhonchi Detection | 1) Modifications related to quantitative measures of performance specifications: Re-training of the ML model with additional data to improve the performance) of the re-trained algorithm compared to the original device while the same type and range of input signal is used<br>2) Modifications related to quantitative measures – technical performance specifications:<br>a. Modification of data preprocessing methodologies<br>b. Data augmentation methodologies<br>c. Architecture and hyper-parameters to improve the performance or the efficiency of the computational resources (running time, memory consumption and CPU utilization).<br>3) Modifications related to device inputs: Expanding the algorithm to include new sources of the same signal type (different model of FDA compatible Stethoscope with equivalent audio acquisition specifications). Modification is limited to expanding to electronic stethoscopes that have FDA 510k clearance (for over-the counter use) at the time that the proposed modification is made. |
| Canvas Dx | 1) Optimization of Decision Thresholds: Adjusting the predefined decision thresholds based on new data without modifying the existing model architecture, parameters, or structure. Optimizing decision thresholds can yield a higher determinate rate while maintaining or improving performance on PPV and NPV. |

| | |
|---|---|
| | 2) Model Retraining: Retraining the model with new data while maintaining the same set of input features and model architecture. Retraining may change feature weights and decision nodes based on the new data. Retraining the model based on new data enables the Device to adapt, improving generalizability and maintaining performance across evolving clinical populations.<br>3) Feature Addition: Adding neurodevelopmental/neurobehavioral features as inputs from the parent, video, or healthcare provider questionnaires, while maintaining the original set of input features and model architecture. Any new features will be limited to those that can be directly observed or reported through existing questionnaires and will not require additional separate testing or assessments. The addition of features may improve Device generalizability and performance, allowing for higher reliability and accuracy of the Device overall. |
| Clarius Prostate AI | 1) Modification of data input sources (Clarius probes) to add data from current Clarius scanners and future 510(K) cleared scanners to the Clarius Prostate AI model so the model can be deployed on more scanners.<br>2) Modification to the filter counts<br>3) Modification of model parameter (initial learning rate) |
| BrightHeart View Classifier | 1) Modification of training and/or validation datasets to update model weights and detection thresholds / Re-training of the BrightHeart View Classifier with new data to optimize its performance.<br>2) Modification of data input sources: ultrasound machine makes (Canon, Fujifilm, SIUI) via retraining of the BrightHeart View Classifier with data from new ultrasound machine<br>3) Addition of standard views (short-axis view at the level of the ventricles and short-axis view at the level of the atrioventricular valves) via retraining with new data from new standard views<br>4) Modification of data input sources: gestational age. Enables the device to achieve sufficient view classification performance to assist users in the identification of standard views for examinations corresponding to gestational ages not previously validated. |
| Clarius Median Nerve AI | 1) Modification of training hyperparameters (initial learning rate, width multiplier, dropout rate)<br>2) Modification of postprocessing algorithms (adjustments to measurement validity thresholds)<br>3) Modification of data input sources (Clarius probes) - To add data from current Clarius scanners and future 510(k) cleared scanners to the Clarius Median Nerve AI model so the model can be deployed on more scanners |

**Supplementary Table 2: Preapproval performance testing described in device approval summaries**

| | Study design | Sample size | Sample demographics | Number of sites | Reference standard | Subgroup analysis |
|---|---|---|---|---|---|---|
| **Caption Guidance** | Prospective | 50 patients | Not described | Not described | Sonographer scans obtained with and without device, graded by expert cardiologists. | Not described |
| | Prospective | 240 patients and 8 device users (RNs) | Not described | Not described | RN scans obtained with device compared to sonographer scans obtained without, graded by expert cardiologists. Expert sonographer | Not described |

| | | | | | | |
|---|---|---|---|---|---|---|
| | | | | | measurements performed on scans obtained with and without device. | |
| **Caption Interpretation Automated Ejection Fraction Software*** | Retrospective | 186 patient studies | Sex, BMI, LVEF values, institution, and ultrasound systems distributions are reported. | Three | LVEF estimation by device compared to biplane method of ejection fraction estimation by cardiologists. Additional metrics compared to expert consensus. | Equivalence across EF, BMI, gender, institution, and ultrasound system (no values reported – they are included in device labelling). |
| **Caption Guidance*** | Not described | Not described | Not described | Not described | Not described | Not described |
| **LINQ II Insertable Cardiac Monitor, Zelda AI ECG Classification System** | Design verification and validation | Not described | Not described | Not described | Not described | Not described |
| **Corvair** | Retrospective | Not described | Not described | Multiple | ECG interpretation of device compared to labelled ECGs in the Common Standards for Quantitative Electrocardiography Standard Database and a proprietary, clinical database. | Not described |
| **Irregular Rhythm Notification Feature*** | Prospective | 573 patients | Age, sex, ethnicity, and race distribution are reported. Mix of Afib history is affirmed. | Not described | Device output compared to a concurrently worn ECG patch | Not described |
| **REMI AI Discrete Detection Module** | Prospective | 50 patients | Age, gender, and monitoring environment (ambulatory or EMU) are reported | Not described | Device output compared to concurrently worn wired EEG, annotated by expert epileptologists | Reported by age and monitoring environment. |
| **Low Ejection Fraction AI-ECG Algorithm** | Retrospective | 16,000 patients | Race, sex, and age are reported. | Four | Compared LVEF calculated by device to echocardiogram obtained within 30 days. | Reported by site, sex, age, race/ ethnicity, BMI, medical history, ECG manufacturer and device. |
| **BoneMRI** | Retrospective | 193 patients | Age, gender, region, and BMI are reported. | Multiple | Compared image quality metrics for BoneMRI scan and concurrently obtained CT scans. | Reported by age, region, and BMI. Equivalence across MRI vendors and field strengths (no values reported). |
| **CLEWICU System** | Retrospective | 11,603 patient stays | Not described | Two | Assessed accuracy of device notification, reference standard not described. | Not described |
| **SleepStageML** | Retrospective | 100 patients | Age distribution is affirmed. Sex and clinical apnea category distributions are reported. | Multiple | Sleep staging of device compared to expert consensus, based on previously acquired polysomnography. | Not described |
| **Clarius OB AI** | Retrospective | 347 patients | Affirm age and ethnic diversity. | 25 | Fetal biometric measurements and gestational age calculation compared | Not described |

| | | | | | | |
|---|---|---|---|---|---|---|
| | | | | | to expert measurement and calculation using Hadlock equations, respectively. | |
| **Acorn 3D Software; Acorn 3DP Model** | Bench testing | Not described | Not described | Not described | Overlap in image segmentation between device and its predicate, accuracy of 3D printed models compared to device-generated digital models | Not described |
| **AiMIFY (1.x)** | Retrospective | 95 patient cases | Affirm T1 input protocol, orientation, acquisition type, scanner, field strength, patient age, sex, pathology, and region diversity (no values reported). | Multiple | Image quality metrics of AiMIFY scans compared to post-contrast MRIs | Not described |
| | Retrospective | 95 patient cases | Same as above | Multiple | Perceived visibility of features and image quality in AiMIFY scans compared to pre- and post-contrast MRIs, rated by neuroradiologists. | Not described |
| **Tyto Insights for Crackles Detection*** | Retrospective | 446 recordings from 445 patients | Age and gender are reported. Dataset obtained via real-world use of device and is indicative of the intended use population. | Not described | Binary device output and probability score compared to pulmonologist consensus and likelihood score, respectively. | Equivalence across age, gender, types of crackles, additional abnormal lung sounds and user type (no values reported). |
| **Sleep Apnea Notification Feature** | Prospective | 1499 patients | Age, sex, race, ethnicity, and sleep apnea severity are reported. Affirm diversity in skin tone and BMI (no values reported). | Multiple | Device output compared to Nox T3s home sleep apnea testing device | Equivalence across all identified subgroups (no values reported). |
| **AI Platform 2.0 (AIP002)** | Retrospective | 100 patients | Affirm diversity in gender, age, race, and ethnicity (no values reported). | Multiple | Device and expert measurements compared. | Not described |
| | Retrospective | 184 patients | Affirm diversity in gender, age, race, ethnicity, device, and pathology (no values reported). | Multiple | Quality rating compared between device and expert sonographers. | Not described |
| | Prospective | 396 scans and 26 users | Affirm range of ultrasound experience among users (no values reported). | Not described | Overall and diagnostic quality rating by expert sonographers | Not described |
| **FETOLY-HEART** | Retrospective | 2288 images and 480 patients | Maternal and gestational age, region, site, BMI, scanner manufacturer, heart abnormality, race, ethnicity, and image quality are reported. | Seven | View and quality criteria detection identified by the device compared to expert consensus | Equivalence across subgroups (no values reported). |
| **Overjet Charting Assist*** | Retrospective | 634 images | Gender, age, and image type are reported | 44 | Device compared to a reference standard established by trained dentists via majority pixel voting. | Reported by gender, region, age, past restorative treatments, dental anatomy, image type, and tooth type. |
| **syngo Dynamics (Version VA41D)** | Retrospective | 150 | Affirm diversity in cardiac condition and representativeness of intended use population. Affirm | Three | Device auto EF calculation compared to Method of Disks (Modified Simpson's Rule) calculated by expert sonographers. | Not described |

| | | | | | | |
|---|---|---|---|---|---|---|
| | | | diversity and report averages for gender, age, and BMI. | | | |
| **HeartFocus (V.1.1.1)** | Retrospective | 290 patients and 361,104 images. | Affirm diversity in BMI, age, and sex (no values reported). | Two | Image quality assessed by the device compared to expert consensus. | Conducted by age, BMI, and sex (no results reported). |
| | Prospective | 240 patients and 8 RNs | BMI, region, cardiac abnormalities, and cardiac implantable devices are reported. Affirm representativeness of intended use population. | Two | Diagnostic and clinical utility of RN scans obtained with device compared to expert scans obtained without, graded by expert cardiologists. | Equivalence across BMI, cardiac abnormalities, scan number in sequence, and study site (no values reported) |
| **Tyto Insights for Rhonchi Detection** | Retrospective | 400 patients | Age and sex reported. Dataset was obtained via real-world use of device so is indicative of the intended use population. | Not described | Binary device output and probability score compared to pulmonologist consensus and likelihood score, respectively. | Equivalence across age, sex, additional abnormal lung sounds and device user (no values reported) |
| **Canvas Dx*** | Prospective | 425 patients | Not described | 14 | Device output compared to expert consensus using DSM-5 criteria | Not described |
| **Clarius Prostate AI** | Retrospective | 139 patients | Affirm diversity in age and ethnicity. | Multiple | Prostate measurements and view identification by device compared to manual expert measurements | Not described |
| **BrightHeart View Classifier** | Retrospective | 2290 images from 579 patients | Not described | Eight | Presence of standard views identified by device compared to expert consensus. | Equivalence across region, ultrasound machine, gestational age, mother's BMI and age, and record type. Worse performance for Asian and Black mothers, possibly due to sample size. No values reported. |
| **Clarius Median Nerve AI** | Retrospective | 182 images from 126 patients | Geographic location is reported. Affirm diversity in ethnicity, gender, and age (no values reported). | Multiple | Device measurements compared to expert consensus. | Not described |

* Data extraction included the predicate device

**Supplemental Table 3: Preapproval human factors testing described in device approval summaries**

| | Sample size | Sample demographics | Number of sites | Subgroup analysis | Outcomes reported |
|---|---|---|---|---|---|
| **Caption Guidance** | Not described | Clinician occupations described (no values reported) | One | Equivalence across clinician occupations affirmed (no values reported). | Critical and non-critical task completion, harmful errors (values not reported). |
| **Caption Guidance*** | 16 users | Prior ultrasound experience of users is reported. | Not described | Not described | Harmful errors, usability (values not reported). |
| **Irregular Rhythm Notification Feature*** | 37 users | Mix of experience with iPhones and Apple Watches affirmed (no values reported). Level of interest in arrhythmia monitoring reported. | Not described | Not described | Safety and efficacy (values not reported), self-reported impact of device outputs on patient decision-making. |

| | | | | | |
|---|---|---|---|---|---|
| **REMI AI Discrete Detection Module** | Not described | Not described | Not described | Not described | Usability of device output evaluated by expert epileptologists (values not reported). |
| **Low Ejection Fraction AI-ECG Algorithm** | Not described | Not described | Not described | Not described | Not described |
| **Clarius OB AI** | Not described | Not described | Not described | Not described | Task completion and usability (values not reported). |
| **Tyto Insights for Crackles Detection*** | Not described | Not described | Not described | Not described | Ability to self-select for the device and risk of self-diagnosis (no values reported). |
| **Sleep Apnea Notification Feature** | Not described | Not described | Not described | Not described | Not described |
| **HeartFocus (V.1.1.1)** | 31 users | Range of user occupations (no values reported). | Not described | Not reported | Critical task completion |
| **Tyto Insights for Rhonchi Detection** | Not described | Not described | Not described | Not reported | Ability to self-select for the device and risk of self-diagnosis (no values reported). |
| **Canvas Dx*** | Not described | Not described | Not described | Not described | Critical task completion, understanding of device use and output (no values reported). |
| **Clarius Prostate AI** | Not described | Not described | Not described | Not described | Task completion (no values reported) |
| **Clarius Median Nerve AI** | Not described | Not described | Not described | Not described | Task completion (no values reported). |

* Data extraction included the predicate device
